# Impact of dosing schedules on performance of rotavirus vaccines in Ghana

**DOI:** 10.1101/2024.06.27.24309591

**Authors:** Ernest O. Asare, Mohammad A. Al-Mamun, George E. Armah, Benjamin A. Lopman, Virginia E. Pitzer

## Abstract

**Background:** Available live-oral rotavirus vaccines are associated with low to moderate performance in low- and middle-income settings. There is limited evidence relating to how the vaccine dosing schedule might be adjusted to improve vaccine performance in these settings.

**Methods:** We used mathematical models fitted to rotavirus surveillance data for children <5 years of age from three different hospitals in Ghana (Korle-Bu Teaching Hospital in Accra, Komfo Anokye Teaching Hospital in Kumasi and War Memorial Hospital in Navrongo) to project the impact of rotavirus vaccination over a 10-year period (April 2012-March 2022). We quantified and compared the impact of the previous vaccination program in Ghana to the model-predicted impact for other vaccine dosing schedules across the three hospitals and the entire country, under different assumptions about vaccine protection. To project the rotavirus vaccine impact over Ghana, we sampled from the range of model parameters for Accra and Navrongo, assuming that these two settings represent the “extremes” of rotavirus epidemiology within Ghana.

**Results:** For the previously implemented 6/10-week monovalent Rotarix vaccine (RV1) schedule, the model-estimated average annual incidence of moderate-to-severe rotavirus-associated gastroenteritis (RVGE) ranged between 1,151 and 3,002 per 100,000 people per year over the 10-year period for the three sites. Compared to no vaccination, the model-estimated median percentage reductions in RVGE ranged from 28-85% and 12-71% among children <1 year and <5 years of age respectively, with the highest and lowest percentage reductions predicted using model parameters estimated for Accra and Navrongo, respectively. The median predicted reductions in RVGE for the whole country ranged from 57-66% and 35-45% among children <1 year and <5 years of age, respectively. The 1/6/10- and 6/10/14-week schedules provided the best and comparable reductions in RVGE compared to the original 6/10-week schedule, whereas there was no improvement in impact for the 10/14-week schedule.

**Conclusions:** We found that administering an additional dose of RV1 might be an effective strategy to improve rotavirus vaccine impact, particularly in settings with low vaccine effectiveness. The results could be extrapolated to other countries using a 2-dose vaccine schedule with low to moderate vaccine performance.

## INTRODUCTION

Routine rotavirus vaccination has been recommended by the World Health Organization (WHO) as the most effective ways to protect infants from rotavirus-associated gastroenteritis (RVGE) morbidity and mortality (1). Since the introduction of rotavirus vaccine, there has been substantial reduction in severe rotavirus diarrhea, rotavirus hospitalizations and rotavirus mortality among children <5 years old (2, 3). However, there is a clear differential in vaccine performance between low- and middle-income countries (LMICs) and high-income countries (HICs), with low to moderate vaccine performance in LMICs (4). In addition to this marked setting-specific variation, there is evidence of within-country variation in rotavirus vaccine performance (5). Thus, there is urgent need to identify novel strategies to improve rotavirus vaccine performance across LMICs.

The monovalent Rotarix vaccine was introduced in the Ghana routine Expanded Program on Immunization in April 2012, with two doses recommended at 6 and 10 weeks of age (6). Ghana subsequently switched to the ROTAVAC vaccine in 2020 with a 3-dose schedule given at 6, 10 and 14 weeks. Despite high vaccination coverage, there has been a varied and modest vaccine impact against RVGE in Ghana compared to HICs (5, 7, 8). Several factors such as co-infections and time of first infection (9–11), malnutrition (12), infant gut microbiome composition (13) and maternal antibodies (14) have been identified as factors explaining the differential rotavirus vaccine performance between HICs and LMICs. Another important factor that could influence vaccine performance is the dosing schedule; however, this has received little attention to date.

Despite the WHO recommendation to relax the age restrictions for rotavirus vaccine due to the low risk of intussusception associated with the vaccine (1), most countries are still using the manufacturer-recommended vaccine dosing schedules (15). This may be partly due to the lack of evidence demonstrating the benefits of changing the dosing schedules for vaccine effectiveness and impact. Despite this, the current flexibility in the dosing schedule provides an opportunity for countries to identify the optimal dosing schedule based on pre- and post-vaccination rotavirus epidemiology. While this is important for countries considering introduction of rotavirus vaccine, it is equally beneficial for countries that want to switch vaccines. However, there has been limited effort to assess the impact of different dosing schedules on the performance of rotavirus vaccines. One study from Ghana showed higher rates of seroconversion following three doses compared to the originally recommended 6 and 10 week schedule, but it unclear how this might correspond to increased vaccine effectiveness and impact (6).

As a complement to clinical trials, mathematical models can be used to investigate the potential impact of dosing schedules on the performance of rotavirus vaccine. However, before these models can be used, it is important to first validate their performance. Several dynamical models have been developed to examine the impact of rotavirus vaccination on morbidity and mortality due to rotavirus (16). One widely-used model for examining the transmission dynamics of rotavirus and impact of vaccination was developed by Pitzer et al. (17). Previous validation of the model showed that it can predict both pre- and post-vaccination rotavirus seasonal patterns and age distributions across different settings, including Ghana (5, 16, 18, 19).

We used our previously validated mathematical model of rotavirus transmission dynamics to quantify the potential impact of changes to the dosing schedules on the performance of rotavirus vaccines in Ghana. We sampled from model parameters estimated from fitting to data from three hospitals in different regions of Ghana separately to project the overall impact of different rotavirus vaccine dosing schedules over 10-year period from April 2012 to March 2022. Our goal was to provide evidence supporting the potential benefits of different dosing schedules for rotavirus vaccine implementation.

## METHODS

### Ethical consideration

The study obtained ethical approval from the Noguchi Memorial Institute for Medical Research of the University of Ghana (Approval Number: CPN:044/12-13) and the Ghana health service ethics review committee (Approval Number: GHS-ERC:06/01/13). A written informed consent was obtained from the parent/guardian of each participant.

### Model description

We used a previously developed age-structured compartmental model of rotavirus transmission dynamics introduced by Pitzer et al. (17), which has been used widely and validated extensively for both pre- and post-vaccination rotavirus transmission dynamics across different settings (5, 16, 18, 19). A detailed description of the model is provided in the supplementary material section 1. In this study, we use previously estimated parameters (Table 1) obtained when the models were fitted separately to pre-vaccination rotavirus inpatient surveillance data from three different hospitals in Ghana (Korle-Bu Teaching Hospital in Accra, Komfo Anokye Teaching Hospital in Kumasi and War Memorial Hospital in Navrongo) (5) to simulate overall rotavirus patterns in Ghana between April 2012 and March 2022. Model vaccine effectiveness estimates (as quantified by the vaccine response rate and duration of vaccine-induced immunity) were based on the observed impact of the Rotarix vaccine introduced in Ghana in April 2012. We previously estimated a significantly higher vaccine response rate and duration of vaccine-induced immunity in Accra compared to Navrongo, with intermediate values estimated for Kumasi (Table 1).

**Table 1.** Previously estimated model parameters obtained from mathematical models fitted to pre- and post-vaccination rotavirus surveillance data from three different hospitals (Accra, Kumasi and Navrongo). The values in parentheses are 95% confidence intervals. See Asare et al. (5) for more details on how the model parameters were estimated. Detailed definitions of the model parameters are provided in supplementary material Table S1. The *S_C1_*, *S_C2_* and *ω_vh_* were estimated assuming heterogeneity in the vaccine response rate (see supplementary material for details).

| Parameter | Definition | Accra | Kumasi | Navrongo |
| --- | --- | --- | --- | --- |
| $R_0$ | Basic reproductive number | 37.861 (36.22-39.54) | 33.661 (29.50-36.20) | 31.529 (30.36-32.78) |
| $1/\omega_m$ | Average duration of maternal immunity (months) | 4.838 (4.46-5.23) | 4.714 (3.46-5.75) | 1.890 (0.39-3.18) |
| $b_1$ | Amplitude of annual seasonal forcing | 0.077 | 0.167 | 0.999 |
| $\phi_1$ | Annual seasonal offset (months) | 7.203 | 4.127 | 1.05 |
| $b_2$ | Amplitude of biannual seasonal forcing | 0.132 | 0.498 | 6.63E-08 |
| $\phi_2$ | Biannual seasonal offset (months) | 1.037 | 1.055 | 5.125 |
| $h$ | Proportion of moderate-to-severe diarrhoea cases reported | 0.107 | 0.017 | 0.012 |
| $d_3$ | Proportion of subsequent infections that are severe | 9.54E-06 | 3.09E-07 | 2.54E-05 |
| $S_C$ | Vaccine response rate (homogeneous response) | 0.989 (0.71-1.00) | 0.649 (0.42-0.88) | 0.203 (0.08-0.32) |
| $1/\omega_v$ | Duration of vaccine-induced immunity assuming homogeneous response (months) | 24.102 (11.11-48.38) | 5.33 (1.23-14.29) | 5.26 (2.46-8.40) |
| $S_{C1}$ | Proportion who responded to the first dose | 0.988 (0.64-1.00) | 0.989 (0.68-1.00) | 0.612 (0.36-0.87) |
| $S_{C2}$ | Proportion who responded to the second dose | 0.878 (0.75-1.00) | 0.892 (0.46-1.00) | 0.19 (0.11-0.48) |
| $1/\omega_{vh}$ | Duration of vaccine-induced immunity assuming heterogeneous vaccine response (months) | 46.451 (12.20-83.33) | 8.194 (2.90-15.36) | 5.465 (3.16-8.33) |

We explored two different scenarios for the vaccine response. For our main analysis, we assumed heterogeneity in vaccine response, in which the probability of “responding” to subsequent vaccine doses (and moving to an immunized compartment in the model) is lower for those who failed to respond to the first dose. As a sensitivity analysis, we assumed homogeneity in vaccine response, in which the probability of responding to each vaccine dose is equal and independent. See supplementary material for details.

### Rotavirus vaccine dosing schedules

We explored various vaccine dosing schedules (Table 2) including current two- and three-dose infant dosing schedules (6/10, 10/14 and 6/10/14 weeks), neonatal dosing schedules (1/6/10 and 1/10/14 weeks) and a booster dosing schedule (6/10/40 weeks). The number of doses and age at which infants receive different doses of the vaccine for each schedule are provided in Table 2.

**Table 2.**
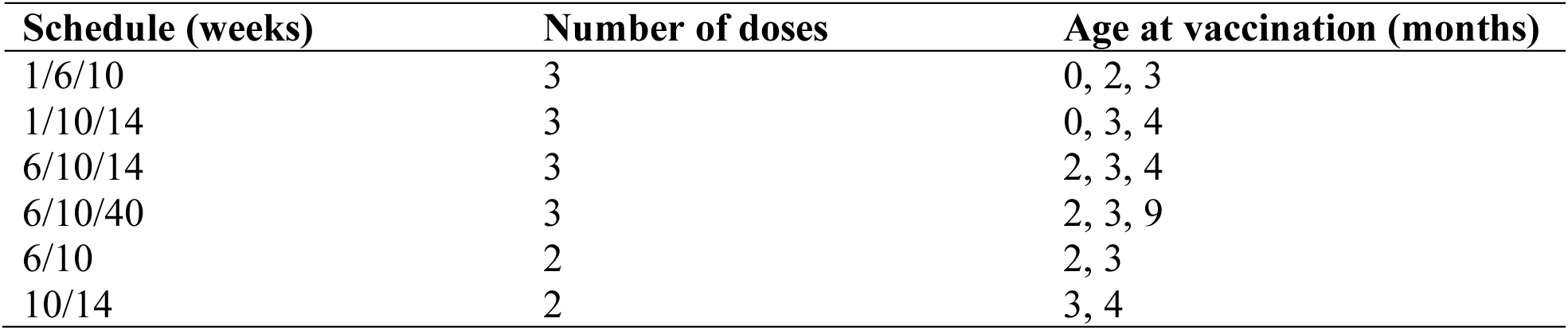
List of considered dosing schedules, the number of doses and age of infants for vaccine administration in the model.

| Schedule (weeks) | Number of doses | Age at vaccination (months) |
| --- | --- | --- |
| 1/6/10 | 3 | 0, 2, 3 |
| 1/10/14 | 3 | 0, 3, 4 |
| 6/10/14 | 3 | 2, 3, 4 |
| 6/10/40 | 3 | 2, 3, 9 |
| 6/10 | 2 | 2, 3 |
| 10/14 | 2 | 3, 4 |

### Simulations of the different dosing schedules

We utilized a beta distribution (20) to sample from the 95% confidence intervals of four key model parameters (transmission rate, duration of maternal immunity, vaccine response rate and duration of vaccine-induced immunity) while using the mean estimates of the other parameters (Table 1). For each setting, 100 parameter sets were generated to predict rotavirus vaccine impact. To simulate rotavirus vaccine impact for the entire country, we sampled from beta distributions of the key model parameters, with the upper and lower bounds defined by the point estimates from Accra and Navrongo, respectively (Table 1).

The overall effect of vaccination for each of the dosing schedules over the 10-year period (April 2012 to March 2022) was calculated as a percentage change using no vaccination as a baseline. The percentage reduction for each of the dosing schedules is given by:

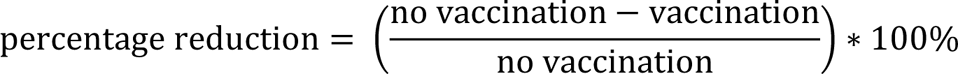

where “vaccination” and “no vaccination” indicate model-estimated moderate-to-severe RVGE cases with and without vaccination, respectively.

## RESULTS

In the absence of rotavirus vaccination, the model estimated the average annual incidence of moderate-to-severe RVGE would range between 2,457 and 3,497 cases per 100,000 person-years over the 10-year period for the three settings (Table 3). The model-projected average annual incidence post-vaccination varied substantially depending on both the vaccine response rate (*S*_C1_ and *S*_C2_) and duration of vaccine-induced immunity (1/*ω_vh_*). For Accra, where we estimated a high vaccine reponse rate and longer duration of vaccine-induced immunity (see Table 1), the projected average annual incidence of RVGE was lowest, ranging from 854 to 1,221 per 100,000 (Table 3). On the other hand, in Navrongo with lower vaccine response rate and shorter duration of immunity (see Table 1), the projected incidence was highest, ranging from 2,743 to 3,003 per 100,000. For Kumasi, the projected incidence was intermediate (ranging from 933 to 1,348 per 100,000) (Table 3). The time series and distribution of model-projected rotavirus infections across the three sites are provided in the supplementary material (see Section 2 for details). The results were similar assuming a homogeneous vaccine response (see supplementary material, Section 3).

**Table 3.** The average and range of model-projected moderate-to-severe RVGE incidence over a 10-year period across the three sites and the whole country (cases per 100,000 person-years).

| Schedule | Accra | Kumasi | Navrongo | Ghana |
| --- | --- | --- | --- | --- |
| Novacc | 2731 (2653-2857) | 2457 (2386-2568) | 3497 (3421-3867) | 3312 (3287-3332) |
| 6/10 | 1151 (920-1694) | 1303 (974-1573) | 3002 (2765-3160) | 2124 (1825-2632) |
| 10/14 | 1221 (1002-1713) | 1348 (1041-1539) | 3003 (2774-3167) | 2123 (1813-2555) |
| 6/10/14 | 874 (519-1679) | 946 (693-1509) | 2743 (2463-2938) | 1797 (1528-2597) |
| 6/10/40 | 930 (589-1693) | 1130 (834-1569) | 2888 (2580-3075) | 1869 (1628-2626) |
| 1/6/10 | 854 (493-1740) | 933 (670-1621) | 2749 (2475-3011) | 1814 (1537-2704) |
| 1/10/14 | 896 (549-1753) | 1100 (818-1684) | 2810 (2515-3027) | 1834 (1565-2709) |

Compared to no vaccination (baseline), the estimated median percentage reduction in moderate-to-severe RVGE among children <5 years varied between dosing schedules and across settings, ranging from 55% to 71% for Accra, 36% to 55% for Kumasi and 12% to 20% for Navrongo over the 10-year period following vaccine implementation (Fig. 1A). Among children <1 year, significantly greater median percent reductions in RVGE were predicted, ranging from 73% to 85% for Accra, 57% to 74% for Kumasi and 28% to 36% for Navrongo (Fig. 1B). The difference in the predicted median percent reduction across settings is lower among children under the age of 1 year (3-fold) than among children under the age of 5 years (6-fold). The estimated reduction differed across schedules, with higher reductions predicted for the 3-dose compared to the 2-dose schedules. The 6/10/14 and 1/6/10 weeks schedules provided the best and comparable vaccine impact, while the lowest and comparable vaccine was predicted for 6/10 and 10/14 weeks schedules. Among the 3-dose schedules, the 6/10/40 schedule had a slightly lower predicted vaccine impact. Results were similar assuming homogeneity in vaccine response, with greater reductions in RVGE incidence for the 3-dose schedules particularly in Accra (see supplementary material, Section 3).

**Fig. 1.**
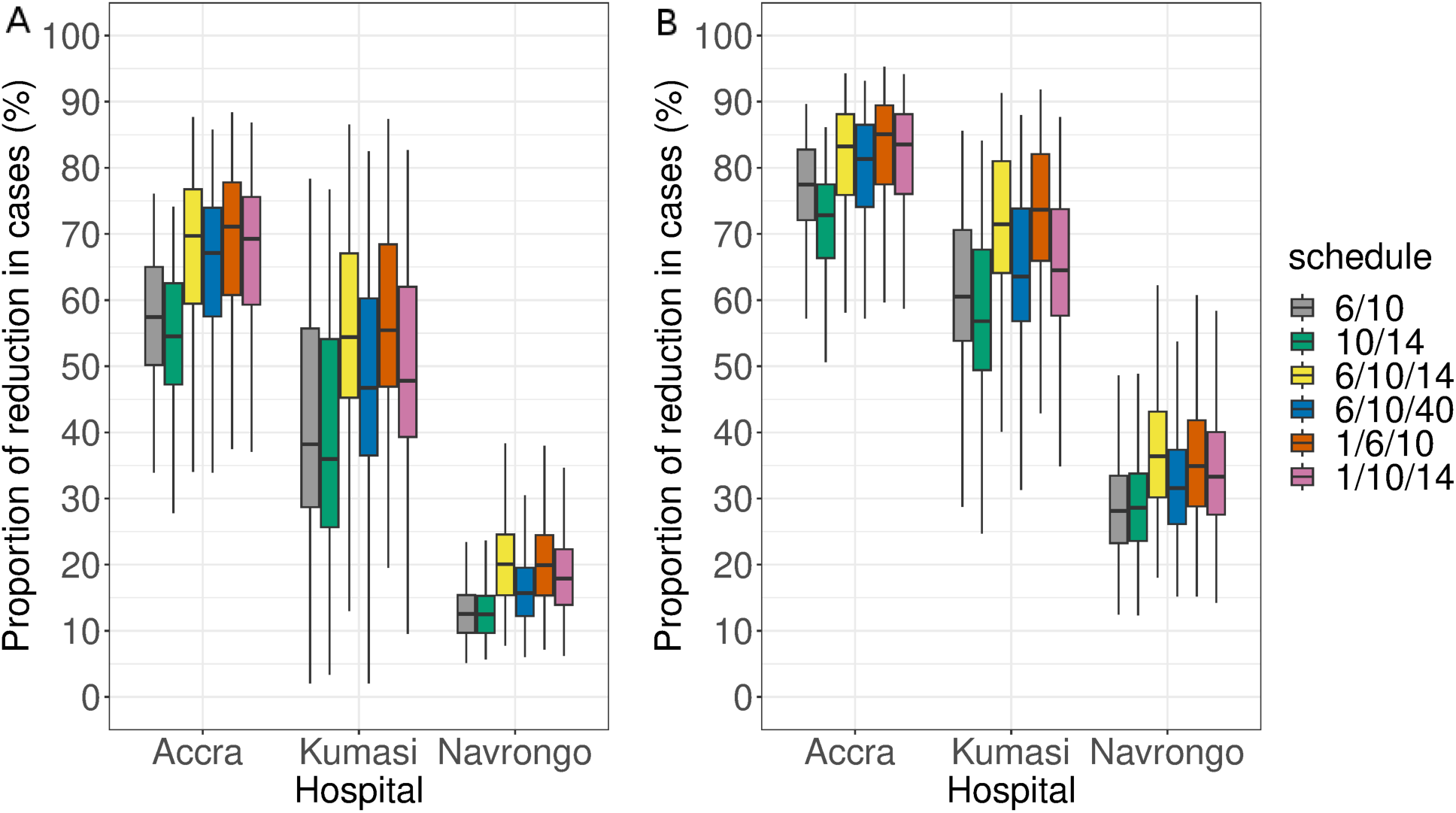
Boxplots of the distribution of the model-projected percentage reductions in rotavirus over the three settings in Ghana between April 2012 and March 2022 using setting-specific estimated model parameters. (A) Children <5 years of age and (B) children <1 year of age. The colours indicate the various dosing schedules.

The results from the overall Ghana scenario (i.e. when we sampled from the full range of estimated model parameters from the three different sites) are shown in Fig. 2. In the absence of vaccination, the highest model-projected number of RVGE cases per month was 928 (<5 years old) and 602 (<1 year old) (Fig. 2A, B). In the absence of vaccination, we estimated that the average annual mean rotavirus incidence over Ghana would have been 3,312 (range 3,287-3,332) per 100,000 over the 10-year period (Table 3). With vaccination, average annual mean rotavirus incidence would range from 1,797 (6/10/14 weeks) to 2,124 (6/10 weeks) per 100,000. The estimated median reduction was highest (45%) and lowest (35%) with the 6/10/14 and 10/14 weeks schedule, respectively (Fig. 2C). Among children <1 year of age, the median percent reductions tended to be higher, ranging between 57% (10/14 weeks) and 66% (6/10/14 weeks) (Fig. 2D).

**Fig. 2.**
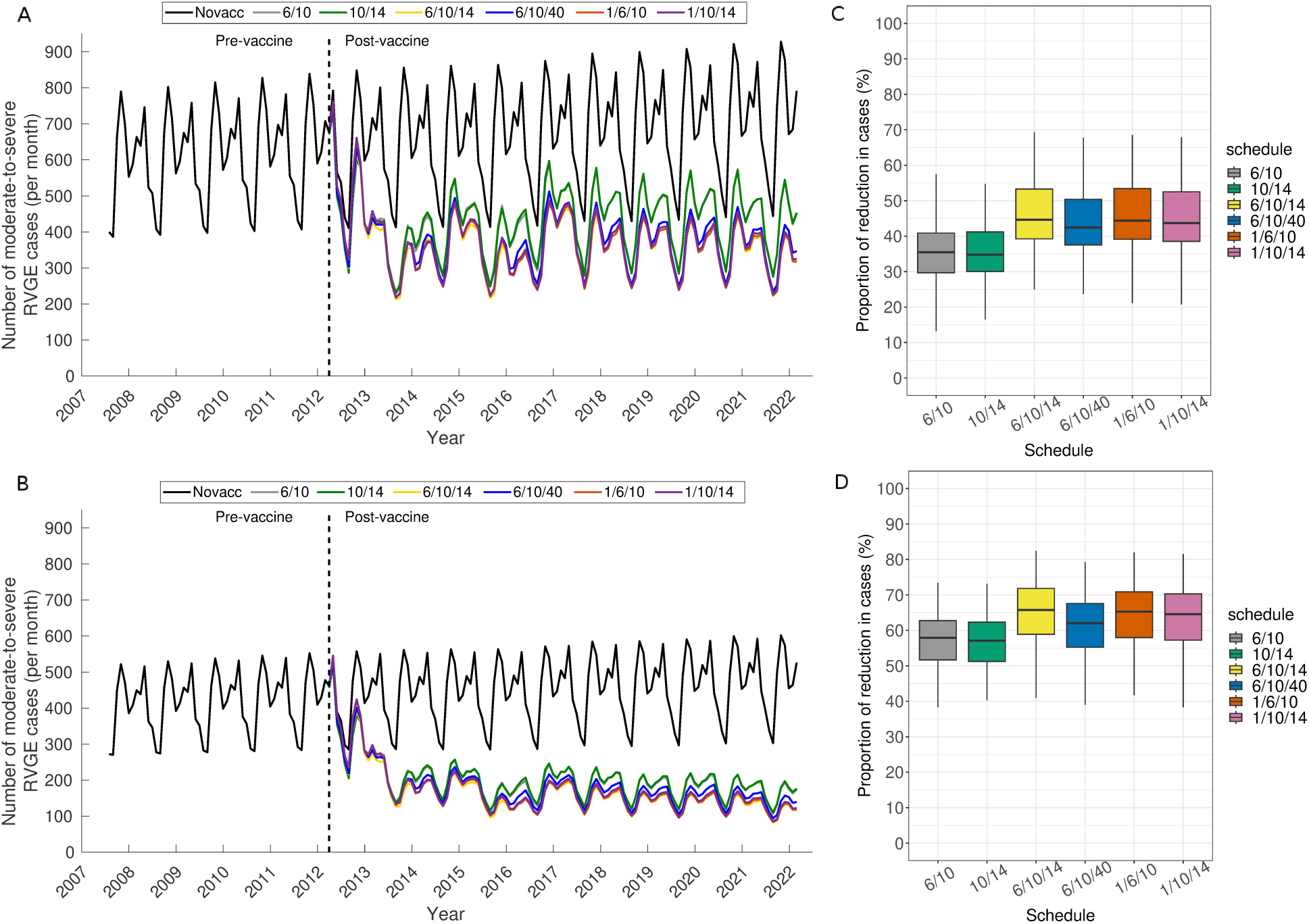
The model projection of rotavirus vaccination over Ghana between April 2012 and March 2022. Time series of average model projected monthly rotavirus cases among children <5 years of age (A) and <1 year of age (B). The lines represent the average from 100 simulations sampled from the range of model-estimated parameters for Accra and Navrongo representing the extremes of rotavirus epidemiology in Ghana. Boxplots of the distribution of the model-projected percentage reductions in RVGE over Ghana for children <5 years of age (C) and <1 year of age (D).

## DISCUSSION

Currently, there is a lack of substantial evidence on the effect of dosing schedules on the potential impact of rotavirus vaccines, resulting in countries still using the manufacturers’ recommended schedules. Our results demonstrate substantial differences in model-projected vaccine impact across different dosing schedules in Ghana, particularly between 3-dose and 2-dose schedules. A 3-dose schedule could provide considerable improvements in vaccine impact compared to the commonly used 2-dose schedule (6/10 and 10/14 weeks) across LMICs. Thus, implementing optimal dosing schedules may substantially improve rotavirus vaccine performance in LMICs.

Despite the modest performance of the vaccine in Navrongo, our projected vaccine impact over Ghana is substantial, ranging from 35-45% (for those under 5 years old) and 57-66% (for those under 1 year old) median reductions in moderate-to-severe RVGE compared to no vaccination. The disparities in vaccine performance across the sites further reveal that studies aimed at evaluating the country-level effectiveness of rotavirus vaccines should consider multiple sites instead of just one in order to provide a more accurate estimate of the vaccine’s impact. Overall, we have shown that the rotavirus vaccine provides substantial health benefits and should be sustained in Ghana.

In the absence of vaccination, the model predicts rotavirus incidence patterns similar to general diarrhea patterns in Ghana (i.e. lower and comparable incidence in Accra and Kumasi and higher incidence in Navrongo) (21). The reasons underlying this may be due to a combination of important factors affecting rotavirus infections, which tend to be favorable in the northern part of the country. Compared to the southern part of the country, the northern part is associated with high prevalence of childhood malnutrition (22), low coverage of water, sanitation and hygiene infrastructure (23) and early exposure to rotavirus infection (24, 25).

Our results reveal substantial differences in the model-projected percentage reduction in RVGE cases across dosing schedules. Compared to no vaccination, 3-dose schedules resulted in a higher percentage reduction in rotavirus cases compared to the 2-dose schedules. This is consistent with data from a randomized clinical trial in Navrongo, Ghana that found a higher seroconversion rate in infants who received 3 doses compared to those who received 2 doses of the Rotarix vaccine (6). However, this higher vaccine response in 3-dose compared with 2-dose recipients is not consistent across LMICs, with some trials showing a higher response rate in 2-dose recipients (26, 27). We also found moderate to substantial variations in the predicted percentage reduction in RVGE among different dosing schedules with the same number of doses, particularly when the duration of vaccine-induced immunity was assumed to be shorter. Thus, both the number and timing of doses administered is important when considering the optimal dosing schedule, which should be carefully selected based on country-specific rotavirus epidemiology.

The highest reductions in RVGE were predicted using parameters estimated from models fitted to the observed vaccine impact in Accra (associated with higher *R*_0_, higher vaccine response rate, and longer duration of vaccine-induced immunity), while the lowest were predicted with estimates from Navrongo (associated with lower *R*_0_, lower vaccine response rate, and shorter duration of vaccine-induced immunity). These findings suggest that alternative or next-generation vaccines with superior effectiveness relative to the current infant rotavirus vaccines are likely to reduce the rotavirus burden across LMICs. However, even with a comparable vaccine response rate between LMICs and HICs, the higher rotavirus transmission rate typically estimated for LMICs compared to HICs (18, 19, 28) could still result in lower vaccine impact in LMICs, but these differences are likely to be minimal compared to what is currently observed. While improvements in sanitation and hygiene practices are likely to reduce the transmission rate of rotavirus, strategies aimed at improving the vaccine response rate in LMICs might have a greater effect in reducing RVGE incidence in these settings.

The timing of vaccination in relation to the age of first rotavirus infection may also play an important role in explaining differences in the predicted reduction in RVGE across the dosing schedules. Several studies have shown that infants who have been infected before vaccination are less likely to seroconvert (9, 10, 25). Thus, the first dose of vaccine needs to be administered early in LMICs before infants are exposed to their first infection to maximize vaccine protection. For instance, a 2-dose schedule of reassortant rotavirus tetravalent vaccine with the first dose administered within 1 month of age provided an efficacy of 63% in Navrongo (25), which is greater than what has been reported for another trial with an infant schedule in Ghana (29). Our results provide some evidence to support this hypothesis, with neonatal schedules (1/6/10 and 1/10/14 weeks) predicted to provide a comparable reduction in RVGE to a 6/10/14 weeks schedule in Navrongo, where we observed earlier infections associated with a shorter estimated duration of maternal immunity (5). While neonatal rotavirus vaccines have yet to be licensed, our results demonstrate that they could offer an improved vaccine performance, as higher vaccine efficacies have been reported for neonatal compared to infant dosing schedules (30, 31). In addition, there is also the potential for an increase in vaccination coverage when the first dose is administered at birth (32).

The interplay between duration of vaccine-induced immunity and the interval between doses can also influence the performance of different dosing schedules. The interval between doses should ideally be shorter than the duration of vaccine-induced immunity. For instance, when the vaccine-induced immunity was assumed to be shorter (5-6 months for Kumasi and Navrongo), the median predicted reduction in RVGE from the booster dose schedule (6/10/40) is substantially lower than the other 3-dose schedules. This could be due to the long interval (∼6 months) between the second and third doses for the 6/10/40 schedule. Using the same model fitted to data from Malawi, a third dose administered at 9 months of age was predicted to provide only a modest improvement in vaccine impact compared to the current 6/10 schedule (18). While a booster or additional doses have been suggested as a strategy for increasing vaccine performance in LMICs, optimal timing for additional doses and the interdose period need to be determined based on the duration of vaccine-induced immunity and rotavirus epidemiology of the country.

An important limitation of this study is that we used the same estimated vaccine response rate and duration of vaccine-induced immunity values obtained when the model was previously fitted to rotavirus surveillance following introduction of a 2-dose monovalent Rotarix vaccine given at 6 and 10 weeks of age in Ghana to evaluate all of the different dosing schedules. The current ongoing neonatal rotavirus vaccine trial in Ghana provides an opportunity to re-evaluate the performance of neonatal dosing schedules against previously (Rotarix) and currently (ROTAVAC) used vaccines in Ghana.

Currently, there is insufficient evidence from randomized clinical trials demonstrating the importance of dosing schedules on rotavirus vaccine performance. Using a mathematical model, we have provided quantitative insights about the potential effect of different dosing schedules on rotavirus vaccine impact. Future research should aim to determine whether it would be cost-effective to include additional vaccine doses considering the increased costs. Given that the WHO has recommended removal of the age restrictions for rotavirus vaccines, it is essential that countries consider alternative dosing schedules and identify the optimal dosing schedule to improve vaccine performance in LMICs. Our model can be a useful tool to identify the optimal country-specific vaccine schedule for countries considering introduction of rotavirus vaccine or switching to a different vaccine.

## Data Availability

All data produced in the present study are available upon reasonable request to the authors

## Supplementary Material

## SECTION 1

### Materials and Methods

The model (Fig. S1) assumes that individuals are born into the maternal antibody protected compartment (*M*) at a rate *B* equal to the national birth rate. This maternally-acquired immunity wanes at rate *ω_m_*, leaving infants susceptible to their primary rotavirus infection (*S*_0_). We assumed that primary infections occur at a rate *λ* and infected individuals (*I*_1_) are infectious for an average duration of 1/*γ*_1_, with only a proportion (*d_1_*) developing severe rotavirus diarrhea. Individuals recover at the end of the infectious period into the *R*_1_ compartment and are assumed to be temporarily immune to reinfection. Immunity wanes at a rate *ω*; after waning of immunity, individuals become susceptible to secondary infection (*S*_1_), which occurs at a reduced rate *σ*_1_*λ*. The secondary infected individuals (*I*_2_) have a lower level of infectiousness (by a factor *ρ*_2_), remain infectious for a shorter duration (1/*γ*_2_), and are less likely to develop severe rotavirus diarrhea (*d*_2_) compared to first infections. Following secondary infection, we assume individuals develop temporary immunity to reinfection (*R*_2_) that wanes at the same rate *ω*. Once this immunity wanes, individuals are transferred into the partially-immune susceptible compartment (*S*_2_) where they become susceptible to subsequent infections that are mostly asymptomatic or mildly symptomatic (*d*_3_) and occur at a reduced rate *σ*_2_*λ*. The subsequent infected individuals (*I*_≥3_) have a further reduced level of infectiousness (by a factor *ρ*_≥3_) and recover at the same rate as the secondary infected individuals (1/*γ*_2_) into the temporary immune compartment (*R*_≥3_). This immunity wanes at the same rate (*ω*), after which individuals re-enter the *S*_2_ compartment.

**Fig. S1.**
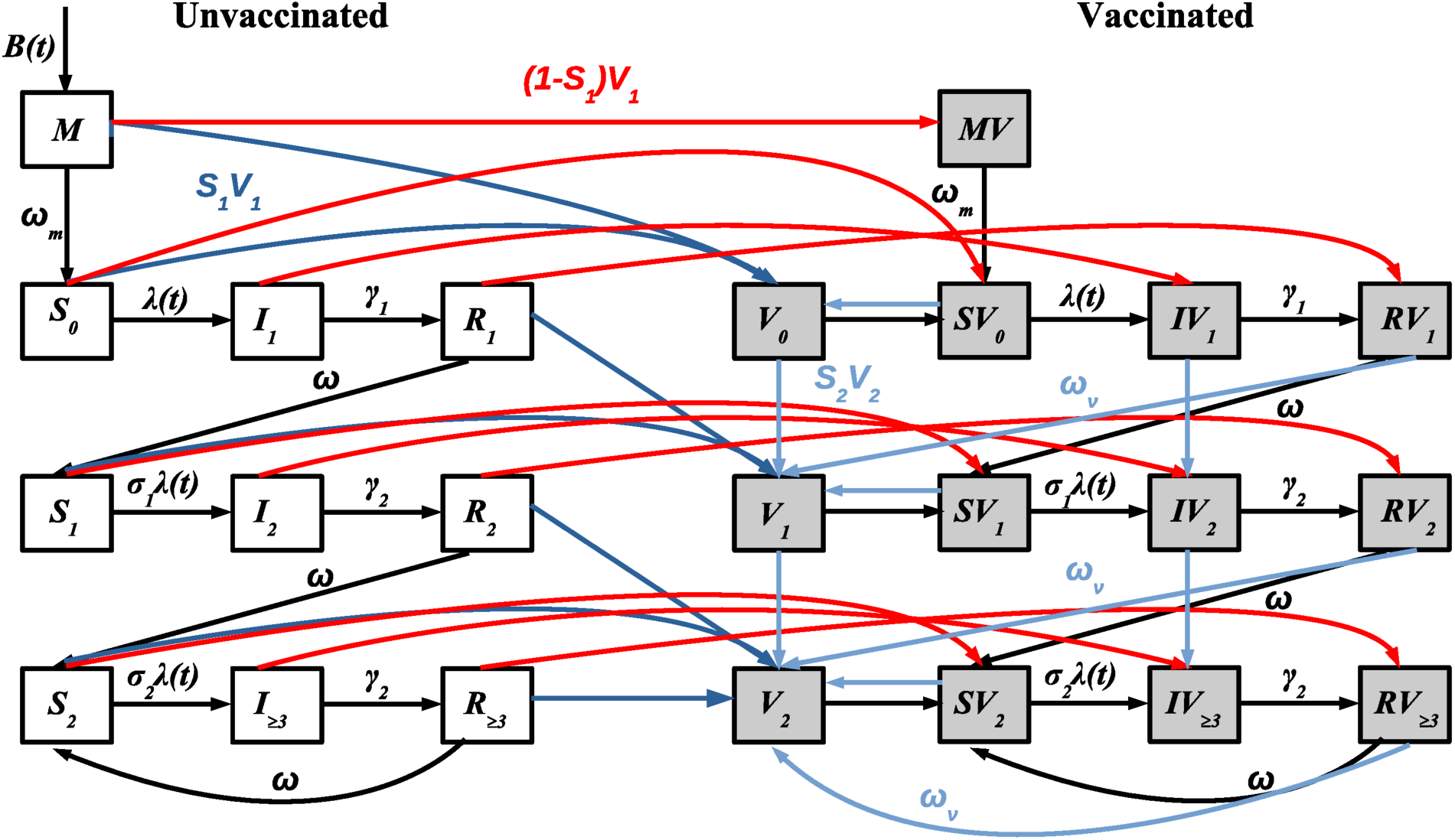
Schematic diagram of the rotavirus transmission model. The open and grey boxes represent vaccinated and unvaccinated individuals, respectively. The *M*, *S*, *I* and *R* represents maternal, susceptible, infected and recovered compartments respectively. The arrows indicate rates of movement among compartments. The red lines represent the transition of individuals who fail to respond to the first dose of rotavirus vaccine. The dark and light blue lines represent the transition of individuals who responded to the first and subsequent doses. The probability of responding to each dose of the vaccine is the same and independent.

The monthly force of infection (rate of transmission from infected to fully susceptible individuals), *λ*(*t*), is given by:

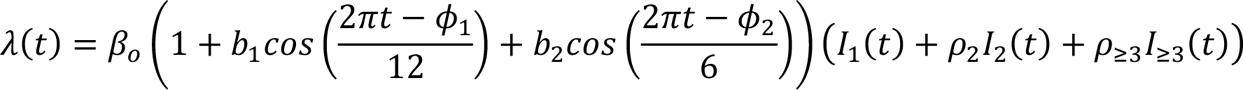

where the model parameters are defined in Table S1.

To incorporate vaccination, we assume that each dose of vaccine confers protection equivalent to one natural infection among those who “respond” to the vaccine dose. Individuals who fail to respond (red lines) to the first dose are transferred to their corresponding state in the vaccinated compartment while those who do not respond to subsequent doses of the vaccine remain in their current vaccinated compartment. For infants who seroconverted (“responded”), vaccination provides temporary immunity against rotavirus infection (*V_i_*) that wanes over time (*⍵_v_*; same for both doses). Following the waning of vaccine-induced immunity, vaccinated infants are transferred to either primary (after first dose) and secondary (after second dose) susceptible compartment, respectively. However, those who responded to both vaccine doses remain protected and are transferred to the next vaccinated-and-protected compartment.

In addition, we refitted our model assuming heterogeneity in the vaccine response rate. Here we assumed that infants who failed to respond to the first dose have lower probability of responding to the second dose (and subsequent doses) compared to those who responded to the first dose. Using previously estimated pre-vaccination model parameters (5), we fitted the model to the post-vaccination data across the three sites while estimating three parameters: proportion of individuals who responded to the first (*S_C1_*) and second vaccine dose (*S_C2_*), and heterogeneous vaccine-derived immunity duration (*ω_vh_*). The probability of responding to the second dose given they failed to respond to the first dose was estimated following Pitzer et al. (18). For the 3-dose schedules, we assumed that the probability of responding to a third dose is the same as the second dose. The age at which infants receive different doses of the vaccine depends on the dosing schedule (see Table 2).

**Table S1.** Model parameters definition and values of fixed parameters. The values of the estimated are provided in Table 1. The *S_C1_*, *S_C2_* and *ω_vh_* were estimated assuming a heterogeneity in vaccine response rate.

| Parameter | Symbol | Value | Reference |
| --- | --- | --- | --- |
| ***Fixed parameters*** |  |  |  |
| Duration of primary infection | $1/\gamma_1$ | 1 week | (33) |
| Duration of secondary infection | $1/\gamma_2$ | 0.5 week | (34, 35) |
| Duration of temporary immunity following infection | $1/\omega$ | 13 weeks | (36) |
| Relative risk of second infection | $\sigma_1$ | 0.62 | (37, 38) |
| Relative risk of third infection | $\sigma_2$ | 0.35 | (37, 38) |
| Relative infectiousness of secondary infection | $\rho_2$ | 0.5 | (39) |
| Relative infectiousness of mild/asymptomatic infections | $\rho_{\geq 3}$ | 0.1 | (39) |
| ***Estimated parameters*** |  |  |  |
| Basic reproductive number | $R_0$ | $=\beta_0/\gamma_1$ | (5) |
| Baseline transmission rate | $\beta_0$ | | (5) |
| Average duration of maternal immunity (months) | $1/\omega_m$ | | (5) |
| Amplitude of annual seasonal forcing | $b_1$ | | (5) |
| Annual seasonal offset (months) | $\phi_1$ | | (5) |
| Amplitude of biannual seasonal forcing | $b_2$ | | (5) |
| Biannual seasonal offset (months) | $\phi_2$ | | (5) |
| Proportion of moderate-to-severe diarrhea cases reported | $h$ | | (5) |
| Proportion of subsequent infections that are severe | $d_3$ | | (5) |
| Vaccine response rate (homogeneous response) | $S_C$ | | (5) |
| Duration of vaccine-induced immunity assuming homogeneous response (months) | $\omega_v$ | | (5) |
| Proportion who responded to the first dose | $S_{C1}$ | Estimated | |
| Proportion who responded to the second dose | $S_{C2}$ | Estimated | |
| Duration of vaccine-induced immunity assuming heterogeneous vaccine response (months) | $\omega_{vh}$ | Estimated | |

## SECTION 2

The annual RVGE incidence for each hospital was calculated by dividing the number of model-predicted moderate-to-severe RVGE cases in a given year by the model-predicted population. The average annual incidence was calculated by dividing the total incidence for each year by 10, the number of years simulated.

Rotavirus transmission patterns vary from strongly seasonal in Navrongo to biannual in Accra (Fig. S2, A-C). In the absence of rotavirus vaccination, the model estimated the average annual incidence of moderate-to-severe rotavirus-associated gastroenteritis (RVGE) would range between 2,457 and 3,312 per 100,000 over the 10-year period for the three settings (April 2012 - March 2022). The impact of vaccination varies by dosing schedule and across the three settings (Fig. S2, A-C). The model-projected average annual incidence of rotavirus over a 10-year period ranges from 854 to 3,003 per 100,000 for Accra and Navrongo, respectively, with vaccination. In the absence of vaccination, the projected proportion of cases in the first year of life is higher for Navrongo (72%) and lower and comparable for Kumasi (59%) and Accra (63%) (Fig. S2, D-F). There is a shift in the proportion of rotavirus cases toward the older age groups following vaccination. This shift varied across dosing schedules and was substantial and comparable from Accra and Kumasi estimates but small from Navrongo estimates (Fig. S2, D-F).

**Fig. S2.**
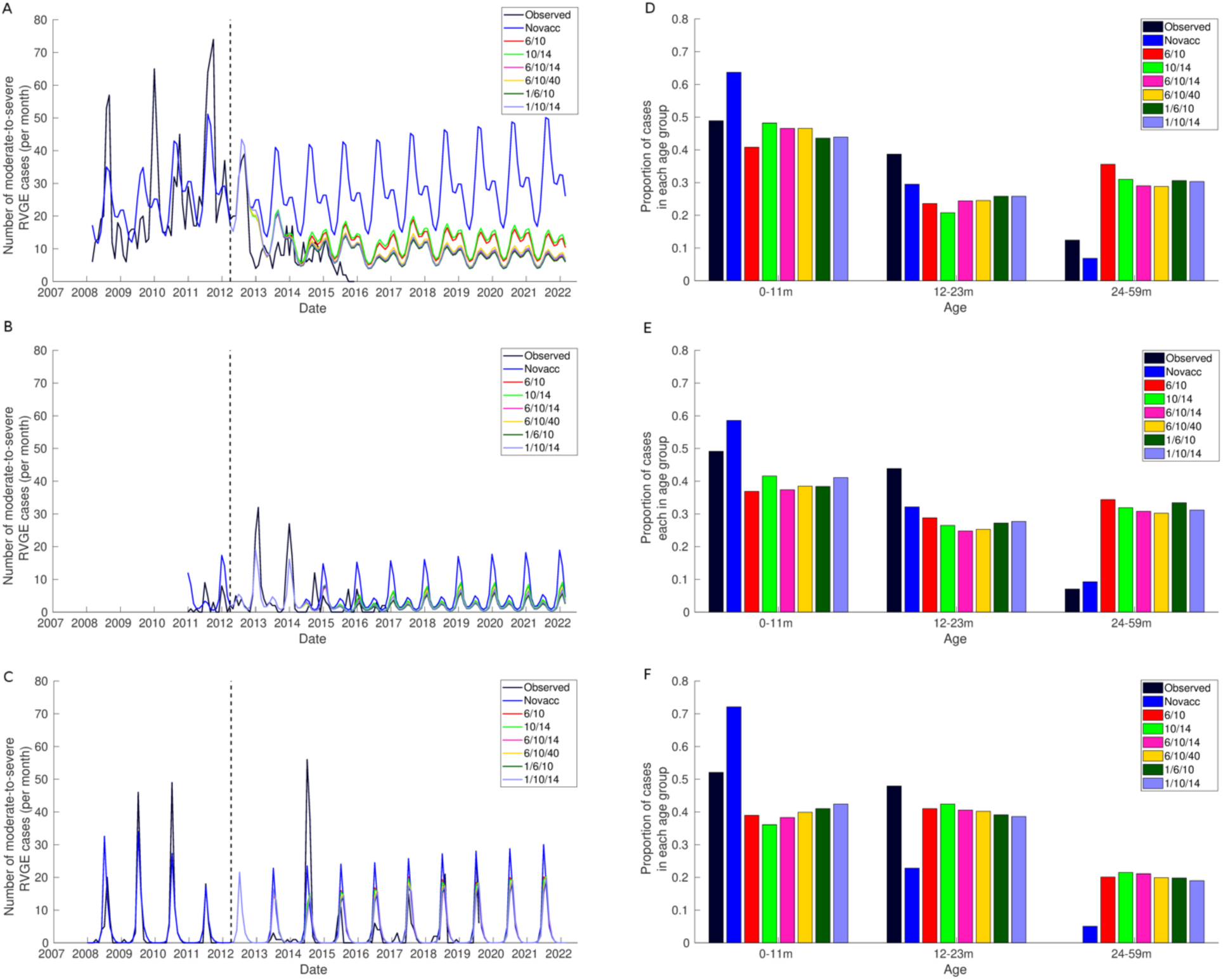
Comparison of laboratory-confirmed and model projected monthly rotavirus cases and age distribution for different dosing schedules across the three settings. (Left) Observed and model-predicted monthly rotavirus cases among children under 5-years for (A) Accra, (B) Kumasi, and (C) Navrongo. The dashed vertical line indicates the date of vaccine introduction (April 2012). The date range of the observed data varied by setting (Accra: August 2007-April 2015; Kumasi: Jan 2009-December 2014; and Navrongo: July 2007-July 2020). (Right) Observed and model-predicted age distribution for (D) Accra, (E) Kumasi, and (F) Navrongo. The age distribution of the model project cases is between April 2012 and March 2022.

## SECTION 3

Results obtained by assuming a homogeneous vaccine response rate are presented here. The results are mostly comparable to those with a heterogeneous vaccine response assumption.

**Fig. S3.**
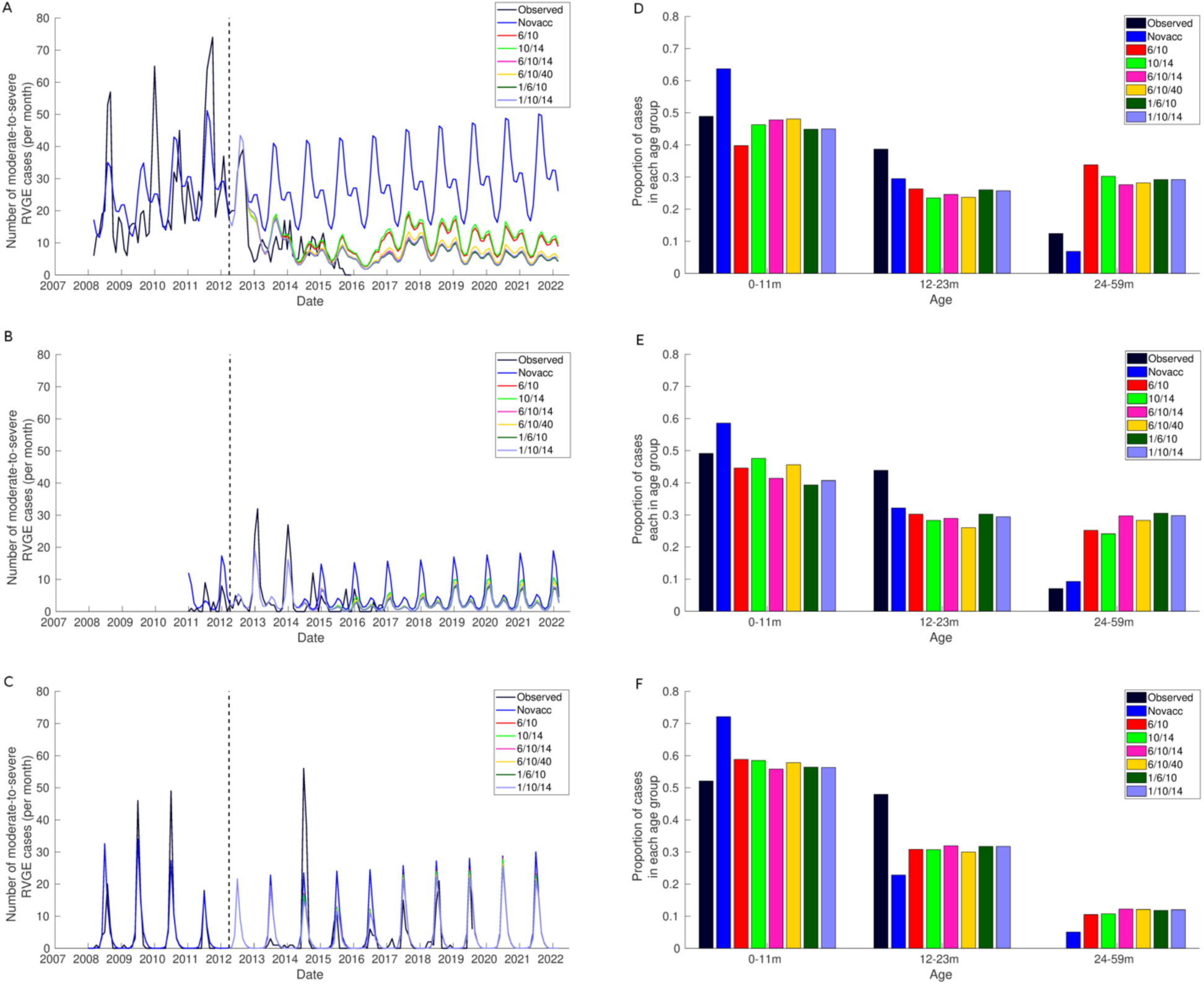
Comparison of laboratory-confirmed and model projected monthly rotavirus cases and age distribution for different dosing schedules across the three settings. (Left) Observed and model-predicted monthly rotavirus cases among children under 5-years for (A) Accra, (B) Kumasi, and (C) Navrongo. The dashed vertical line indicates the date of vaccine introduction (April 2012). The date range of the observed data varied by setting (Accra: August 2007-April 2015; Kumasi: Jan 2009-December 2014; and Navrongo: July 2007-July 2020). (Right) Observed and model-predicted age distribution for (D) Accra, (E) Kumasi, and (F) Navrongo. The age distribution of the model project cases is between April 2012 and March 2022.

**Fig. S4.**
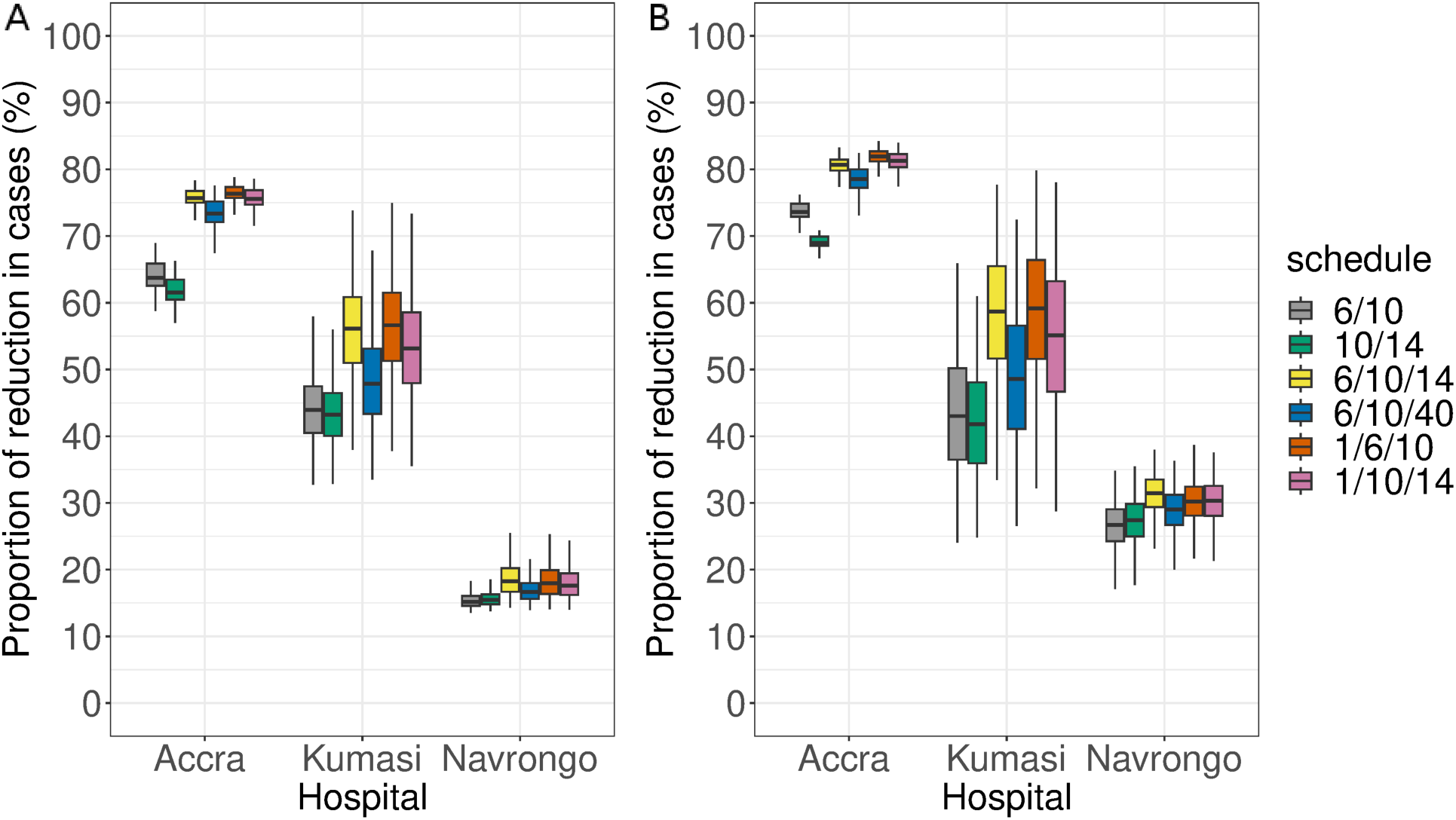
Boxplots of the distribution of the model-projected percentage reductions in rotavirus over the three settings in Ghana between April 2012 and March 2022 using setting-specific estimated model parameters. (A) Children <5 years of age and (B) children <1 year of age. The colours indicate the various dosing schedules.

**Fig. S5.**
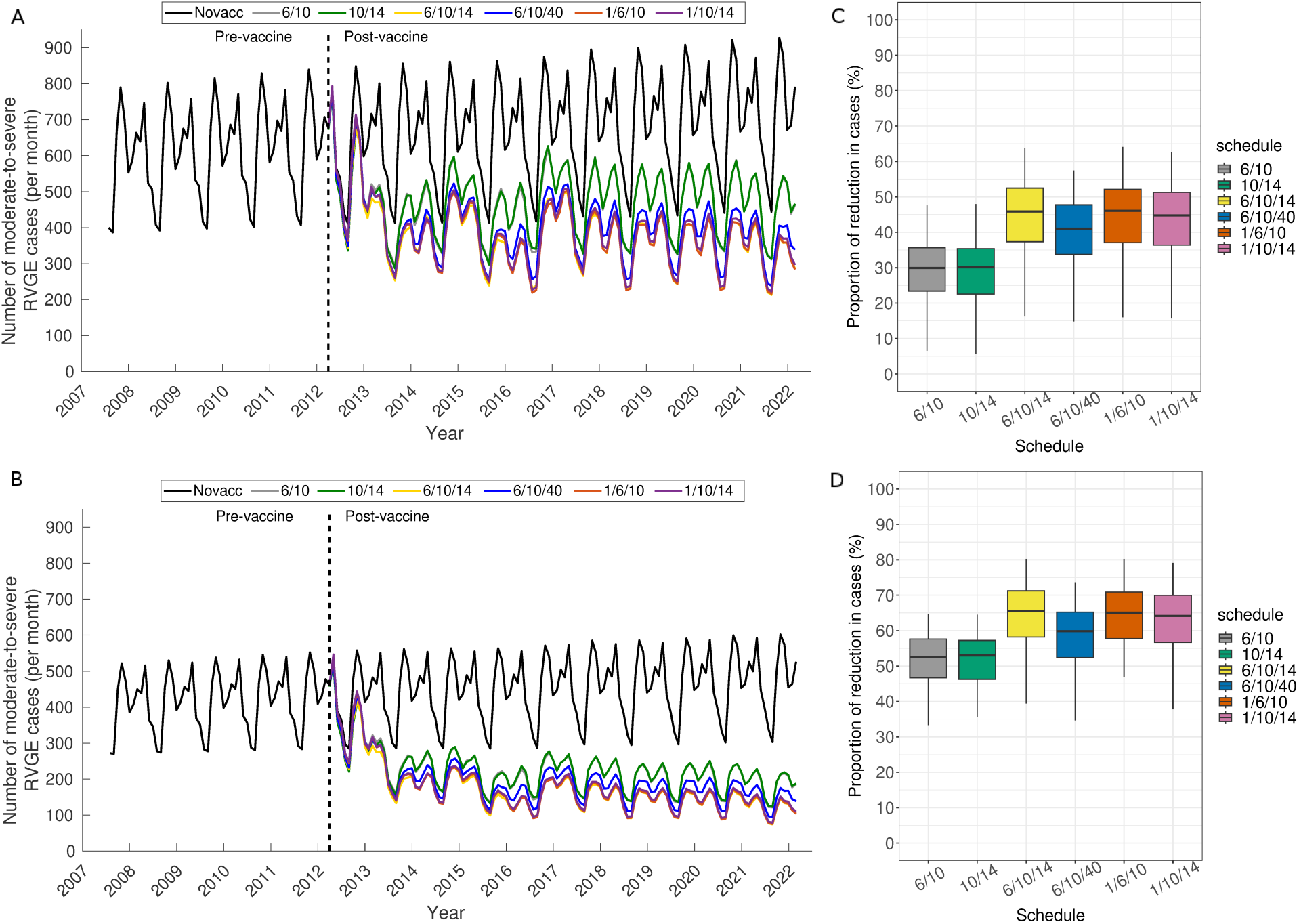
The model projection of rotavirus vaccination over Ghana between April 2012 and March 2022 using the range of values sampled from estimated model parameters for Accra and Navrongo assuming a homogeneous vaccine response rate. Time series of average model projected monthly rotavirus cases among children <5 years of age (A) and <1 year of age (B). The lines represent the average from 100 simulations sampled from point estimates of the model estimated parameters for Accra and Navrongo representing the extremes of rotavirus epidemiology in Ghana. Boxplots of the distribution of the model-projected percentage reductions in RVGE over Ghana for children <5 years of age (D) and <1 year of age (E).

